# Examination of Medication Use Patterns by Age Group, Comorbidity, and Month in COVID-19 Positive Patients in a Large State-wide Health System During the Pandemic in 2020

**DOI:** 10.1101/2021.07.15.21259539

**Authors:** Jonathan H. Watanabe, Jimmy Kwon, Bin Nan, Shira R. Abeles, Sanjay R. Mehta

**Author notes:** (Corresponding Author) Jonathan H. Watanabe, PharmD, PhD.

## Abstract

**Objectives:** Main objective was to systematically determine most frequently used medications among COVID-19 patients overall and by hospitalization status. Secondary objective was to measure use patterns of medications considered potential therapeutic options

**Design:** Retrospective cohort study.

**Setting:** The five academic medical centers of University of California Health.

**Participants:** University of California COVID Research Data Set (UC CORDS) patients between March 10, 2020 and December 31, 2020.

**Exposure(s):** Confirmed COVID-19 positive by SARS-CoV-2 nucleic acid amplification.

**Main Outcome(s) and Measure(s):** Main outcomes were percentages of patients prescribed medications, overall, by age group, and by comorbidity based on hospitalization status. Use percentage by month of COVID-19 diagnosis was measured. Cumulative count of potential therapeutic options was measured over time.

**Results:** Dataset included 22897 unique patients with COVID-19 (mean [SD] age, 42.4 [20.4] years; 12154 [53%] female). Among the sample, 6326 28%) were non-Hispanic White, 8475 (37%) were Hispanic, 1562 (7%) Asian, and 1313 (6%) Black. A COVID-related hospitalization occurred in 3546 patients. Of the hospitalized patients, more than 30% had baseline comorbidities of hypertension (48%), hyperlipidemia (37%), and type 2 diabetes (35%). Most frequently used medications in patients overall were acetaminophen (21.2%), albuterol (14.9%), ondansetron (13.9%), and enoxaparin (10.8%). Medications used were generally similar across ages and comorbidities. Prior to May, dexamethasone was rarely used, with well under 50 COVID-19 patients that had been hospitalized to that point receiving the medication. By mid-August, more than 500 patients to that point had received dexamethasone. Hydroxychloroquine use effectively halted in COVID-19 hospitalized patients after May. Throughout the period of March to December 2020, enoxaparin was used in the most patients to that point at any instance. By mid-December, more than 2000 in the analysis cohort of hospitalized patients had received enoxaparin.

**Conclusions and Relevance:** In this retrospective cohort study, across age and comorbidity groups, predominant utilization was for supportive care therapy. Dexamethasone and remdesivir experienced large increases in use. Conversely, hydroxychloroquine and azithromycin use markedly dropped. Medication utilization rapidly shifted towards more evidence-concordant treatment of patients with COVID-19 as rigorous study findings emerged.

## Introduction

Coronavirus disease 2019 (COVID-19) is a pandemic infection caused by the novel severe acute respiratory syndrome coronavirus 2 (SARS-CoV-2). Awareness of the SARS-CoV-2 infectious capacity coupled with a case fatality rate greater than 0.6%^1^ has led to global efforts to reduce transmission and identify effective treatment approaches. As of July 9, 2021 there have been more than 185 million confirmed cases and in excess of 4 million deaths worldwide.^2^

Understanding medication use patterns for patients with confirmed COVID-19 will provide needed insight into the evolution of COVID-19 treatment over the course of the SARS-CoV-2 Pandemic and provide evidence for clinical management considerations. The goal for this analysis was to characterize the frequency of medications used based on percentage of COVID-19 positive individuals receiving the medications overall, by age group, and by comorbid condition for both hospitalized and non-hospitalized patients. To examine how this pattern has changed since the pandemic began, medication use based on month of diagnosis was tracked. To quantify use of medications considered as possible treatments that impact the clinical course by shortening the duration of symptoms or preventing complications (referred hereafter as ‘potential therapeutic options’), dexamethasone, remdesivir, enoxaparin, heparin, colchicine, hydrocortisone, tocilizumab, azithromycin, hydroxychloroquine and medication classes of angiontensin-2 converting enzyme inhibitors (ACEIs) and angiotensin receptor blockers (ARBs), ^3–9^ number of hospitalized patients in the cohort that used each PTO were counted over the pandemic.

## Methods

The University of California COVID Research Data Set (UC CORDS) was constructed to be a timely data set for research purposes distributed weekly, containing SARS-CoV-2 testing results and COVID-19 treatment information collected from across University of California (UC) Health. It is a Health Insurance Portability and Accountability Act (HIPAA) Limited Data Set generated from all five UC Health medical centers (Davis, Irvine, Los Angeles, San Diego, San Francisco). In 2019, UC Health had 1,092,522 inpatient days. This was equivalent to 6.6% of the total inpatient days in the state. Care was provided to 1.8 million unique patients by UC Health in 2019 which is 4.6% of the population of California that year. ^10–12^ UC CORDS was operationalized by UC Health as ‘non-human subjects research’ and analyses are considered institutional review board exempt. The Strengthening the Reporting of Observational Studies in Epidemiology (STROBE) reporting guideline was followed for this analysis.^13^

Data for 22897 patients confirmed positive with COVID-19 by SARS-CoV-2 RNA nucleic acid amplification with probe detection was extracted for the period of March 10, 2020 through December 31, 2020. Consistent with UC Health reporting, patients were categorized as experiencing a COVID-19-related hospitalization if they were admitted to the hospital within 30 days of confirmed test or tested positive during their hospitalization.

### Generation of Comorbidities from EHRs

Patients with COVID-19 with comorbid conditions of hypertension, hyperlipidemia, type 2 diabetes, COPD/Asthma, heart disease, chronic kidney disease (CKD), cancer, obesity, the respiratory diseases asthma and COPD, and those receiving dialysis are at increased risk for poor outcomes.^14–16^ To examine medication utilization in patients diagnosed with COVID-19 with these risk factors at baseline, we first determined the presence of these conditions using the *International Classification of Diseases, Tenth Revision, Clinical Modification* (ICD-10-CM) codes in the patient electronic health record (EHR). Given the possibility that preexisting conditions would not be reported at COVID-19 diagnosis, ICD-10-CM codes up to one-year preceding diagnosis were included. Self-reported gender and race/ethnicity were extracted from EHR as well as body mass index (BMI) for hospitalized patients. Age was estimated based on patient birth year as birthdate was removed from UC CORDS for de-identification. Uses of mechanical ventilation and extracorporeal membrane oxygen (ECMO) were extracted for each hospitalized patient using affiliated *International Classification of Diseases, Tenth Revision, Procedure Coding System* codes and Current Procedural Terminology, 4^th^ Edition codes.

Medications used for each patient were determined from the EHR by measurement of all medication active ingredients utilized within 30 days of COVID-19 positive test based on the RxNorm standardized nomenclature for clinical drugs from the National Library of Medicine.^17^ Given the assumption, in context of COVID-19, of a similar therapeutic class effect of angiotensin converting enzyme-2 inhibitors (ACEIs) and angiotensin receptor blockers (ARBs) usage of these medications was collapsed to the overall category of ‘ACEIs/ARBs’. Monthly utilization was based on month in which patient was confirmed for COVID-19.

### Statistical Analysis

Descriptive measurement of demographic and clinical characteristics was performed for COVID-19 positive patients in the sample overall and by hospitalization status. Continuous data were expressed as mean +/-SD or median and interquartile range [IQR] values. Use was quantified by percentage of patients with corresponding 95% confidence interval. All confidence intervals and subgroup sample sizes are included in the affiliated tables.

Data were extracted with module pyodbc, version 4.0.30 in Python, version 3.8.3 (Python Software Foundation). All analyses performed in R, version 3.6.3 (R Project for Statistical Computing). Statistical significance defined using two-sided significance level of alpha = 0.05.

## Results

### Descriptive Statistics

The dataset contained 22897 unique patients confirmed positive for COVID-19 between March 10, 2020 and December 31, 2020 (mean [SD] age, 42.4 [20.4] years; 12154 [53%] female; median [25^th^, 75^th^ percentile] BMI for hospitalized patients, 28.5 [23.8, 32.9]. Among the sample, 6326 (28%) were non-Hispanic White, 8475 (37%) were Hispanic, 850 (7%) Asian, 738 (6%) Black and 6013 (47%) reported as ‘other’. Of the 22897 unique COVID-19 test confirmed patients, 2584 patients had a COVID-related hospitalization. Of the hospitalized patients, the most common comorbidity was hypertension (48%). Greater than 20% of hospitalized patients had hyperlipidemia (37%), type 2 diabetes (35%), heart disease (24%), and CKD (22%). The rates of Asthma/COPD and cancer in hospitalized patients were 17% and 13%, respectively (Table 1).

**Table 1.** Descriptive Characteristics of the COVID-19 Test Confirmed Patients

|  | Tested COVID-19 Positive n(%) | Non-Hospitalized n(%) | Hospitalized n(%) |
| --- | --- | --- | --- |
| **Age** |  |  |  |
| Sample Size | 22897 | 19351 | 3546 |
| mean (sd) | 42.35 ± 20.40 | 39.92 ± 19.35 | 55.63 ± 20.84 |
| under 50 | 14,379 (63) | 13,121 (68) | 1,258 (35) |
| 50 to <65 | 4,985 (22) | 4,014 (21) | 971 (27) |
| 65 or older | 3,533 (15) | 2,216 (11) | 1,317 (37) |
| **Gender** |  |  |  |
| Female | 12,154 (53) | 10,573 (55) | 1,581 (45) |
| **Race & Ethnicity** |  |  |  |
| non-Hispanic White | 6,326 (28) | 5,417 (28) | 910 (26) |
| Hispanic | 8,475 (37) | 6,768 (35) | 1,707 (48) |
| Black | 1,313 (6) | 1,041 (5) | 272 (8) |
| Asian | 1,562 (7) | 1,262 (7) | 300 (8) |
| Native Hawaiian / Other Pacific Islander / American Indian / Alaska Native | 252 (1) | 200 (1) | 52 (1) |
| Other | 3,100 (14) | 2,378 (12) | 722 (20) |
| Not Reported | 7,012 (31) | 6,316 (33) | 696 (20) |
| **Body Mass Index of Hospitalized Patients** |  |  |  |
| median (25th, 75th) | NA | NA | 28.5 (23.8, 32.9) |
| < 18.5 | NA | NA | 98 (3) |
| 18.5 to < 25.0 | NA | NA | 849 (26) |
| 25 to < 30.0 | NA | NA | 1,004 (30) |
| 30 or more | NA | NA | 1,364 (38) |
| **Variable** |  |  |  |
| Hypertension | 4,200 (18) | 2,503 (13) | 1,697 (48) |
| Hyperlipidemia | 3,721 (16) | 2,411 (12) | 1,310 (37) |
| Type 2 Diabetes | 2,685 (12) | 1,427 (7) | 1,258 (35) |
| Asthma/COPD | 1,795 (8) | 1,198 (6) | 597 (17) |
| Heart Disease | 1,520 (7) | 670 (3) | 850 (24) |
| Chronic Kidney Disease | 1,326 (6) | 548 (3) | 778 (22) |
| Cancer | 1,394 (6) | 921 (5) | 473 (13) |
| Baseline Dialysis | 190 (1) | 46 (0) | 144 (4) |
| **Hospital Procedures** |  |  |  |
| Mechanical Ventilation | 661 (3) | 0 (0) | 661 (19) |
| ECMO | 54 (0) | 0 (0) | 54 (2) |
| **Positive Covid Test Month** |  |  |  |
| March | 591 (3) | 478 (2) | 113 (3) |
| April | 921 (4) | 699 (4) | 222 (6) |
| May | 709 (3) | 467 (2) | 242 (7) |
| June | 1,915 (8) | 1,542 (8) | 373 (11) |
| July | 3,789 (17) | 3,202 (17) | 587 (17) |
| August | 2,027 (9) | 1,617 (8) | 410 (12) |
| September | 1,108 (5) | 863 (4) | 245 (7) |
| October | 1,083 (5) | 880 (5) | 203 (6) |
| November | 3,200 (14) | 2,872 (15) | 328 (9) |
| December | 7,554 (33) | 6,731 (35) | 823 (23) |

### Medication utilization overall and for patients by hospitalization status by age group, comorbidity, hospital procedure status

#### All Patients that Tested Positive for COVID-19

The ten medications used in the largest percentage of confirmed COVID-19 patients over the study period overall were acetaminophen (21.2%), albuterol (14.9%), ondansetron (13.9%), enoxaparin (10.8%), sennosides (9.3%), lidocaine (8.5%), polyethylene glycol (PEG) 3350 (8.4%), dexamethasone 8.1%, azithromycin (7%), insulin lispro (6.8%). Acetaminophen, ondansetron, albuterol, enoxaparin, and sennosides were part of the top five medications for every age category except for those under 50 years old where lidocaine was present rather than sennosides (Table 2). Across comorbidities, medications were consistent with those observed in the age group analysis. For any of the baseline comorbidities, the medication used in the largest proportion was acetaminophen. Usage of acetaminophen ranged from 39.6% in patients with Cancer to 61.5% for patients with CKD. Enoxaparin or heparin was a top ten medication for all comorbidities. For patients that received dialysis at baseline, most frequent medications were acetaminophen, heparin, insulin lispro, ondansetron, lidocaine, sennosides, albuterol, pantoprazole, vancomycin, and PEG 3350 (Table 3).

**Table 2.** Medication Utilization by Age Group

All Positive COVID-19 Patients
| Rank | Overall (n = 22896) | Percent | 95% CI | <50 (n = 14379) | Percent | 95% CI | 50 to <65 (n = 4984) | Percent | 95% CI | 65 or older (n = 3533) | Percent | 95% CI |
| --- | --- | --- | --- | --- | --- | --- | --- | --- | --- | --- | --- | --- |
| 1 | acetaminophen | 21.2 | ( 20.6, 21.7) | acetaminophen | 14.3 | ( 13.7, 14.9) | acetaminophen | 26.2 | ( 25.0, 27.4) | acetaminophen | 41.8 | ( 40.2, 43.4) |
| 2 | albuterol | 14.9 | ( 14.4, 15.3) | albuterol | 9.4 | ( 8.9, 9.9) | albuterol | 20.7 | ( 19.6, 21.8) | albuterol | 28.8 | ( 27.3, 30.3) |
| 3 | ondansetron | 13.9 | ( 13.5, 14.4) | ondansetron | 9.4 | ( 8.9, 9.9) | ondansetron | 17.6 | ( 16.5, 18.7) | ondansetron | 27.1 | ( 25.6, 28.6) |
| 4 | enoxaparin | 10.8 | ( 10.4, 11.2) | enoxaparin | 5.6 | ( 5.2, 6.0) | enoxaparin | 15.5 | ( 14.5, 16.5) | sennosides, USP | 25.8 | ( 24.4, 27.2) |
| 5 | sennosides, USP | 9.3 | ( 8.9, 9.7) | lidocaine | 5.4 | ( 5.0, 5.8) | sennosides, USP | 12.4 | ( 11.5, 13.3) | enoxaparin | 25.3 | ( 23.9, 26.7) |
| 6 | lidocaine | 8.5 | ( 8.1, 8.9) | ibuprofen | 5.0 | ( 4.6, 5.4) | dexamethasone | 11.7 | ( 10.8, 12.6) | polyethylene glycol 3350 | 23.2 | ( 21.8, 24.6) |
| 7 | polyethylene glycol 3350 | 8.4 | ( 8.1, 8.8) | sennosides, USP | 4.2 | ( 3.9, 4.5) | ACEIs/ARBs | 11.2 | ( 10.3, 12.1) | atorvastatin | 21.3 | ( 19.9, 22.7) |
| 8 | dexamethasone | 8.1 | ( 7.8, 8.5) | polyethylene glycol 3350 | 4.0 | ( 3.7, 4.3) | insulin lispro | 11.0 | ( 10.1, 11.9) | dexamethasone | 20.3 | ( 19.0, 21.6) |
| 9 | azithromycin | 7.0 | ( 6.6, 7.3) | azithromycin | 4.0 | ( 3.7, 4.3) | polyethylene glycol 3350 | 10.8 | ( 9.9, 11.7) | ACEIs/ARBs | 19.1 | ( 17.8, 20.4) |
| 10 | insulin lispro | 6.8 | ( 6.5, 7.1) | benzonatate | 4.0 | ( 3.7, 4.3) | benzonatate | 10.7 | ( 9.8, 11.6) | heparin | 19.0 | ( 17.7, 20.3) |

Hospitalized Patients
| Rank | Overall(n = 3545) | Percent | 95% CI | <50(n = 1258) | Percent | 95% CI | 50 to <65(n = 970) | Percent | 95% CI | 65 or older(n = 1317) | Percent | 95% CI |
| --- | --- | --- | --- | --- | --- | --- | --- | --- | --- | --- | --- | --- |
| 1 | acetaminophen | 88.6 | ( 87.6, 89.7) | acetaminophen | 86.8 | ( 84.9, 88.7) | acetaminophen | 89.6 | ( 87.7, 91.5) | acetaminophen | 89.6 | ( 87.9, 91.3) |
| 2 | enoxaparin | 62.1 | ( 60.5, 63.7) | ondansetron | 61.9 | ( 59.2, 64.6) | enoxaparin | 70.6 | ( 67.7, 73.5) | sennosides, USP | 63.0 | ( 60.4, 65.6) |
| 3 | ondansetron | 59.0 | ( 57.4, 60.6) | enoxaparin | 56.6 | ( 53.9, 59.3) | albuterol | 62.2 | ( 59.1, 65.3) | enoxaparin | 61.2 | ( 58.6, 63.8) |
| 4 | sennosides, USP | 54.1 | ( 52.4, 55.7) | sennosides, USP | 43.2 | ( 40.5, 45.9) | ondansetron | 59.2 | ( 56.1, 62.3) | albuterol | 58.2 | ( 55.5, 60.9) |
| 5 | albuterol | 53.3 | ( 51.7, 55.0) | lidocaine | 42.8 | ( 40.1, 45.5) | sennosides, USP | 56.1 | ( 53.0, 59.2) | polyethylene glycol 3350 | 57.5 | ( 54.8, 60.2) |
| 6 | polyethylene glycol 3350 | 49.2 | ( 47.5, 50.8) | albuterol | 41.3 | ( 38.6, 44.0) | polyethylene glycol 3350 | 49.0 | ( 45.9, 52.1) | ondansetron | 56.0 | ( 53.3, 58.7) |
| 7 | lidocaine | 42.5 | ( 40.8, 44.1) | polyethylene glycol 3350 | 40.7 | ( 38.0, 43.4) | insulin lispro | 47.7 | ( 44.6, 50.8) | heparin | 47.8 | ( 45.1, 50.5) |
| 8 | dexamethasone | 40.1 | ( 38.5, 41.7) | docusate | 35.1 | ( 32.5, 37.7) | dexamethasone | 44.4 | ( 41.3, 47.5) | dexamethasone | 45.9 | ( 43.2, 48.6) |
| 9 | heparin | 38.8 | ( 37.2, 40.4) | fentanyl | 30.8 | ( 28.2, 33.4) | lidocaine | 40.0 | ( 36.9, 43.1) | insulin lispro | 45.6 | ( 42.9, 48.3) |
| 10 | insulin lispro | 38.4 | ( 36.8, 40.1) | dexamethasone | 30.6 | ( 28.1, 33.1) | heparin | 39.1 | ( 36.0, 42.2) | lidocaine | 43.9 | ( 41.2, 46.6) |

Non-Hospitalized Patients
| Rank | Overall(n = 19351) | Percent | 95% CI | <50(n = 13121) | Percent | 95% CI | 50 to <65(n = 4014) | Percent | 95% CI | 65 or older(n = 2216) | Percent | 95% CI |
| --- | --- | --- | --- | --- | --- | --- | --- | --- | --- | --- | --- | --- |
| 1 | acetaminophen | 8.8 | ( 8.4, 9.2) | acetaminophen | 7.4 | ( 7.0, 7.8) | acetaminophen | 10.9 | ( 9.9, 11.9) | acetaminophen | 13.4 | ( 12.0, 14.8) |
| 2 | albuterol | 7.8 | ( 7.5, 8.2) | albuterol | 6.4 | ( 6.0, 6.8) | albuterol | 10.7 | ( 9.7, 11.7) | ACEIs/ARBs | 11.6 | ( 10.3, 12.9) |
| 3 | ondansetron | 5.7 | ( 5.3, 6.0) | ondansetron | 4.4 | ( 4.0, 4.8) | ondansetron | 7.5 | ( 6.7, 8.3) | albuterol | 11.4 | ( 10.1, 12.7) |
| 4 | azithromycin | 3.7 | ( 3.4, 4.0) | ibuprofen | 3.0 | ( 2.7, 3.3) | ACEIs/ARBs | 7.3 | ( 6.5, 8.1) | ondansetron | 9.9 | ( 8.7, 11.1) |
| 5 | ACEIs/ARBs | 3.6 | ( 3.3, 3.8) | azithromycin | 2.7 | ( 2.4, 3.0) | azithromycin | 5.6 | ( 4.9, 6.3) | atorvastatin | 9.5 | ( 8.3, 10.7) |
| 6 | benzonatate | 3.1 | ( 2.9, 3.4) | benzonatate | 2.3 | ( 2.0, 2.6) | benzonatate | 5.1 | ( 4.4, 5.8) | azithromycin | 6.4 | ( 5.4, 7.4) |
| 7 | ibuprofen | 2.8 | ( 2.6, 3.0) | fluticasone | 2.2 | ( 1.9, 2.5) | atorvastatin | 5.1 | ( 4.4, 5.8) | amlodipine | 5.2 | ( 4.3, 6.1) |
| 8 | fluticasone | 2.7 | ( 2.5, 2.9) | lidocaine | 1.8 | ( 1.6, 2.0) | metformin | 4.6 | ( 4.0, 5.2) | dexamethasone | 5.1 | ( 4.2, 6.0) |
| 9 | atorvastatin | 2.5 | ( 2.3, 2.7) | guaifenesin | 1.6 | ( 1.4, 1.8) | fluticasone | 3.9 | ( 3.3, 4.5) | aspirin | 5.1 | ( 4.2, 6.0) |
| 10 | guaifenesin | 2.3 | ( 2.1, 2.5) | codeine | 1.6 | ( 1.4, 1.8) | guaifenesin | 3.8 | ( 3.2, 4.4) | metoprolol | 5.0 | ( 4.1, 5.9) |

**Table 3.**
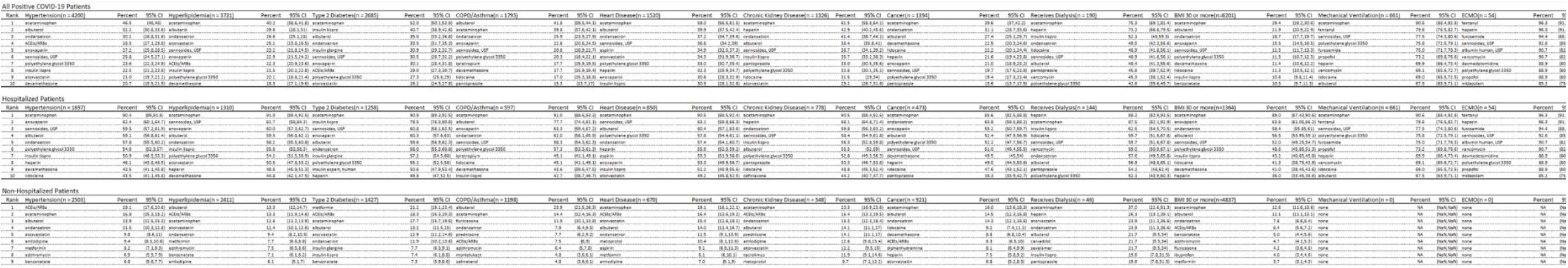
Medication Utilization by Baseline Comorbidity and Hospital Procedure

#### Hospitalized Patients with COVID-19

The ten medications used in the largest percentage of confirmed COVID-19 hospitalized patients were acetaminophen (88.6%), enoxaparin (62.1%), ondansetron (59%), sennosides (54.1%), albuterol (53.3%), PEG 3350 (49.2%), lidocaine (42.5%), dexamethasone (40.1%), heparin (38.8%), insulin lispro (38.4%), Medications were similar across age groups. Dexamethasone was among the top ten medications for all age groups (Table 2). Heparin was among the top ten for all comorbidities except for COPD/Asthma. For patients that received dialysis at baseline, top medications were heparin, acetaminophen, insulin lispro, lidocaine, sennosides, vancomycin, ondansetron, albuterol, pantoprazole, and PEG 3350. For obese patients, the medications in the top ten were consistent with the overall list for hospitalized patients (Table 3).

For hospitalized COVID-19 patients that had mechanical ventilation, top medications were acetaminophen, fentanyl, sennosides, PEG 3350, furosemide, propofol, heparin, vancomycin, lidocaine, and albuterol. For patients that received ECMO, top medications were fentanyl, heparin, furosemide, sennosides, albumin, vancomycin, dexmedetomidine, PEG 3350, propofol, and midazolam (Table 3).

#### Non-Hospitalized Patients with COVID-19

The proportion of individuals receiving each medication was substantially lower in non-hospitalized patients. The ten medications used in the most patients were acetaminophen (8.8%), albuterol (7.8%), ondansetron (5.7%), azithromycin (3.7%), ACEIs/ARBs (3.6%), benzonatate (3.2%), ibuprofen (2.8%), fluticasone (2.7%), atorvastatin (2.5%), and guaifenesin (2.3%). Among all age groups, acetaminophen was the most common ranging from 7.4% in those under 50 years old to 13.4% for those 65 years old and above. Albuterol and ondansetron were among the top ten for all age groups (Table 2). For all comorbidities studied, ACEIs/ARBS were among the six most commonly used medications and were the first or second most common for hypertension (19.1%), hyperlipidemia (13.3%), type 2 diabetes (18.3%), heart disease (16.4%), and CKD (16.4%). Ondansetron was among the ten most commonly used medications for all comorbidities. For patients that had baseline dialysis, top medications were acetaminophen, heparin, atorvastatin, ondansetron, albuterol, carvedilol, sevelamer, insulin lispro, pantoprazole, albumin (Table 3).

### Medication utilization by month of COVID-19 diagnosis

#### All patients that tested positive for COVID-19

In March, among all patients COVID-19 test confirmed, the most common medications were acetaminophen (21.3%), albuterol (20.5%), azithromycin (16.2%), ondansetron (12.5%), guaifenesin (11.7%), enoxaparin (11.2%), PEG 3350 (10.5%), hydroxychloroquine (10.2%), ceftriaxone (9.6%), and sennosides (9.6%). By April, while the most common medications were similar, use of azithromycin and hydroxychloroquine dropped. Consequently, March was the only month in which hydroxychloroquine appeared among the top ten. April was the last month in which ceftriaxone was among the top ten. Enoxaparin was the sixth most commonly used medication in March at 11.2%, but increased in May to the third most used medication at 22.8%. Enoxaparin remained in the top five medications through December.

Dexamethasone was observed as a top ten medication for the first time in July (8.3% of patients), increasing to 11.1% in August, and remained among the top eight prescribed meds thereafter. Heparin was not among the top ten medications after September (Table 4).

**Table 4.**
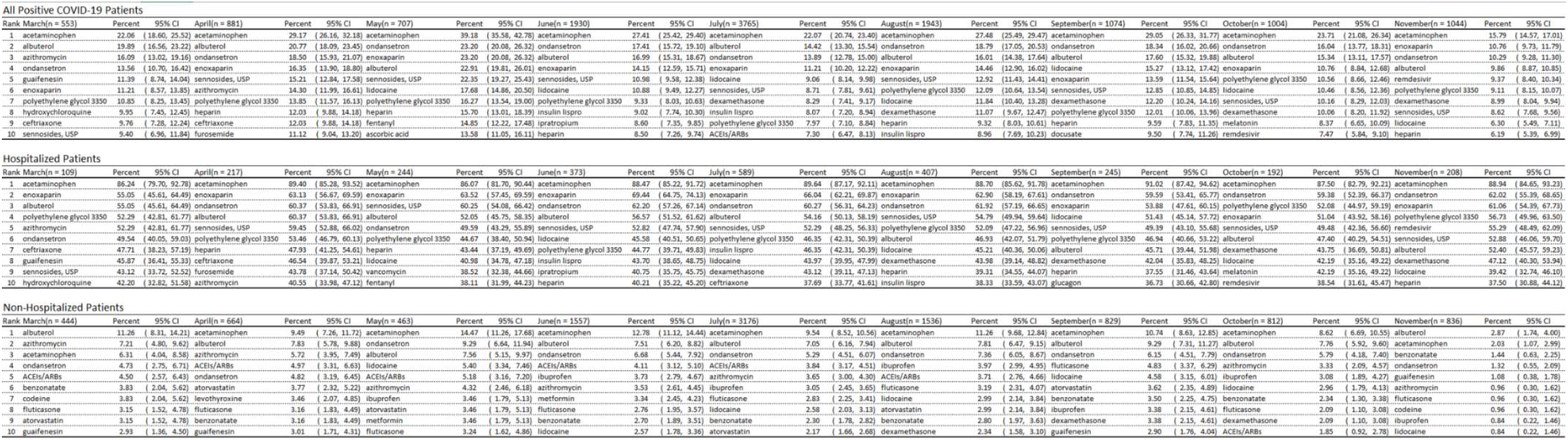
Medication Utilization by Month

#### Hospitalized Patients with COVID-19

In March, the most common medications for those hospitalized were acetaminophen (85.8%), enoxaparin (56.6%), albuterol 56.6%, azithromycin (54%), PEG 3350 (51.3%), ondansetron (48.7%), ceftriaxone (48.7%), guaifenesin (47.8%), sennosides (45.1%), and hydroxychloroquine (43.4%). The top two medications were acetaminophen and either enoxaparin or ondansetron for every month. From March to April, azithromycin use dropped from 54% to 40.5%. Hydroxychloroquine declined rapidly from a high of 43.4% in March and was not in the ten most common medications for the duration of the study. Early in the Pandemic, ceftriaxone was frequently used (48.7% and 45.9% of patients in March and April, respectively). After April, however, ceftriaxone was no longer found among the ten most frequently used medications. Heparin was not among the top ten medications after September. Dexamethasone was used in 43.3% of patients in July with usage exceeding 50% in November and December. Remdesivir, approved under FDA emergency use authorization in May 2020, was among the top ten medications in October at 39.9% and increased to 52.7% in December (Table 4).

#### Non-Hospitalized Patients with COVID-19

In March, the top medications were albuterol (11.9%), azithromycin (7.3%), acetaminophen (6.1%), ACEIs/ARBs (4.8%), ondansetron (4%) codeine (4%), benzonatate (3.8%), guaifenesin (3.1%), fluticasone (2.9%), atorvastatin (2.9%). While azithromycin use decreased after March, it remained among the top ten in all months except August and September. Albuterol and acetaminophen were among the top three medications throughout. Fluticasone was among the top ten medications in all months besides October and December. Dexamethasone was among the top ten medications from August through December (Table 4).

### Cumulative Use Count in Hospitalized Patients Since March 2020 of Potential Treatment Option Medications in Hospitalized Patients

Throughout the period of March to December 2020, enoxaparin was used in the most patients to that point at any instance. By mid-December, more than 2000 in the analysis cohort of hospitalized patients had received enoxaparin. Prior to May, dexamethasone was rarely used, with well under 50 COVID-19 patients that had been hospitalized to that point receiving the medication. By mid-August, more than 500 patients to that point had received dexamethasone. By mid-December, dexamethasone use increased to the extent that more than 1000 patients to that point had received it. Hydroxychloroquine use effectively halted in COVID-19 hospitalized patients after May. Azithromycin prescribing slowed from initial heavy use. Among the PTOs studied, it was the third most frequently used in patients as of August 1, but by close of 2020 had fallen to the sixth most commonly used. Increase in counts of heparin, ACEIs/ARBs, and hydrocortisone use was fairly stable over time. Total dexamethasone users exceeded azithromycin users by end of August. Roughly 1000 patients had received remdesivir by close of December (Figure 1).

**Figure 1.**
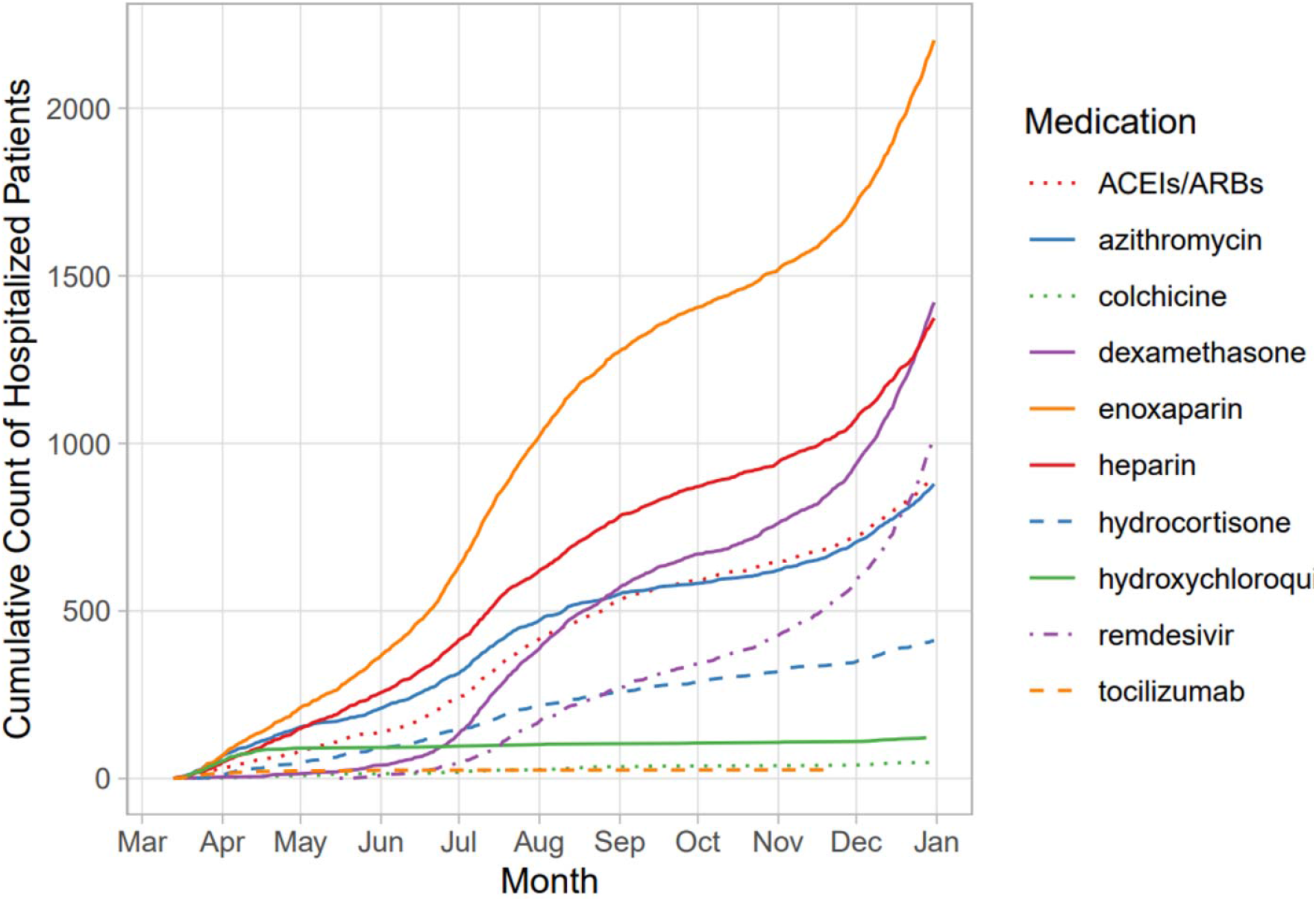
Cumulative Count of Hospitalized Patients Using Potential Therapeutic Option Over Time

## Discussion

This study is the first comprehensive analysis of medication utilization for COVID-19 patients in a large, diverse, state-wide health system overall and by comorbidity, age group, baseline dialysis use, and by month for both the hospitalized and non-hospitalized. Numbers of patients that used medications considered potential therapeutic options during the Pandemic were also tracked over time.

Initially, in the Pandemic, our data shows attempts were made to use antimicrobials to manage COVID-19. Azithromycin, ceftriaxone, and hydroxychloroquine were each used in more than 40% of hospitalized patients in March. This quickly abated. By May, none of the three were amongst the top ten medications for hospitalized patients. Supportive care medications for symptom relief of fever, nausea, pain, cough, constipation, and shortness of breath were heavily used throughout with acetaminophen being used in the largest percentage of all COVID-19 patients followed by albuterol, ondansetron, enoxaparin, and sennosides. Enoxaparin had over 50% use throughout 2020 as enoxaparin serves both as supportive care for deep vein thrombosis prophylaxis and for treatment, given the thrombophilia caused by COVID-19.^19^ Examining other potential therapeutic option over time, dexamethasone and remdesivir use increased substantially. Use of remdesivir likely corresponded with availability, as early in the Pandemic, it was only available through trials. On the other hand, hydroxychloroquine use dropped from over 40% in late March and was not observed in top ten medications beyond that month. Early studies of limited sampled size favored use of these agents,^20,21^ but later larger, controlled studies did not demonstrate clinical benefit in patients.^7,22^

Treatment for COVID-19 patients has progressed from the initial stages, where treatment decisions were educated based on case series and in vitro data. With little guidance, the early approaches applied by clinicians were best guesses. Remarkably, practice patterns appeared to change in near real-time as quality data became available through landmark trials. The trajectories of medication usage generally corresponded to publication of rigorous studies. The acceleration in use of dexamethasone is reasonably correlated to the publication of findings from the RECOVERY trial.^3^ The wide accessibility of data, effectively at the moment of publication, to clinicians over the course of the Pandemic is unparalleled compared to past public health crises. This was demonstrated by the potential therapeutic option curves where usage was generally concordant with the emerging evidence over time. A prior study examined outpatient prescriptions for hydroxychloroquine noted a decline after May 2020. However, this analysis was not focused on patients confirmed for COVID-19.^23^ An analysis over a two month period that examined medication use patterns in COVID-19 hospitalized patients also showed reductions in azithromycin and hydroxychloroquine over time.^24^ Differentiating strengths of our study compared to these prior studies was a longer duration (March to December 2020) to observe oscillations in use of these agents. Beyond a small selection of medications, we captured use patterns of the most common medications prescribed by age, high-risk condition, and month.

Overall, this analysis also demonstrated that most of the medications used to treat COVID-19 patients have been generally inexpensive, generic medications. While this is comforting, more expensive proprietary medications have now been released under EUA (e.g. monoclonal antibodies). Furthermore, generic medications have been increasingly subject to shortages. This compilation of medication use from the COVID-19 experience can serve as a guide for medication needs during future pandemics from respiratory viruses.

The population of COVID-19 patients in the CORDS features a demographic breakdown consistent overall with the State of California. Hispanics were 37% of the study population which is similar to 39% of the state. Non-Hispanic Whites were 28% of COVID-19 positive patients in this study population compared to 37% in the state. Percentages of those in the sample that reported to be Black, and Native Hawaiian/Pacific Islander/American Indian/Alaska Native were similar to the demographic breakdown of California (7% and 1%, respectively).^12^

### Limitations

Outpatient medications for patients in CORDS may require at least a month for complete capture. Use rates are conservative estimates for the non-hospitalized patients since medications for COVID-19 positive patients prescribed medications outside of UC Health are not present in CORDS. While the study demographics were consistent with California overall, given, that this state has a large minority population this may influence generalizability to the US. Consequently, the percentage of COVID-19 positive cases that were Non-Hispanic White in CORDS (28%) was lower than that observed in US national estimates (56%). Hispanics were 37% of the positive cases in our dataset and 21% in the US. A smaller percentage of COVID-19 positive patients were black (6%) than observed in national estimates (12.2%). Asians were 7% of the COVID-19 positive patients in CORDS compared to 4% in US estimates.^25^

## Conclusions

In this retrospective cohort study, across age and comorbidity groups, predominant utilization was for supportive care therapy. Dexamethasone and remdesivir experienced large increases in utilization over time. Conversely, hydroxychloroquine and azithromycin use rapidly declined. Anticoagulation was recognized early in the Pandemic as an important approach for both prophylaxis and in treatment and was widely applied throughout. Medication utilization has changed, in near real-time, in the direction of evidence-concordant treatment of patients with COVID-19 as quality data became available.

## Data Availability

The extracted dataset used for this retrospective analysis is maintained on secured servers and is only available for analysis via virtual desktop maintained and supported by the University of California Irvine Health that manages access to the University of California COVID-19 Research Data Set (UC CORDS).

